# Methylphenidate for Children and Adolescents with Attention Deficit Hyperactivity Disorder (ADHD) – unpublished or unregistered randomised clinical trials

**DOI:** 10.1101/2023.08.24.23293874

**Authors:** Magnus Tang Kristensen, Christian Gluud, Ole Jakob Storebø

## Abstract

**Introduction:** This is a follow-up study on our recent systematic review by Storebø and colleagues on methylphenidate for children and adolescents with attention-deficit/hyperactivity disorder (ADHD) published in The Cochrane Library. We aim to investigate the risks of research waste and publication bias in randomised clinical trials investigating methylphenidate versus placebo or no intervention for children and adolescents with ADHD.

**Method:** The method used includes our initial cohort of randomised clinical trials selected from searching Clinicaltrials.gov and the EUCTR with the following criteria: methylphenidate versus placebo or no intervention with or without co-interventions for children or adolescents with ADHD, randomised clinical trials, any dosage, any delivery method, and at least 75% children and adolescents with ages less than 18 years as well as a time period of 1999-2022.

Our primary objective is to assess how many randomised clinical trials of methylphenidate on children and adolescents with ADHD are registered in protocol databases, but never published in academic literature or as tabular summary results. The number of participants included in these trials is a secondary objective. Our third objective is to examine the relationship between trial results and the chance of being published. Our fourth objective is to assess the time from registry to publication of randomised clinical trials of methylphenidate on children and adolescents with ADHD in either a journal or as summary results, and the number of participants in these trials. The cutoff time for a publication to be considered timely published will be 6 months, as per European Medicines Agency guidelines for clinical trials involving children.

**Results:** This is our protocol for the study and no results are presently available.

## Background

This is a follow-up study on a recent systematic review by Storebø and colleagues [1]. This review suggested potential benefits of methylphenidate on ADHD core symptom severity as well as general behavior, but the certainty of the evidence for the use of methylphenidate for ADHD in children and adolescents was found to be low or very low in all 212 randomised clinical trials [1]. Furthermore, most trials were quite small, with the average sample size being only 70 children. This calls into question small trial effects and publication bias, which is commonly analysed via funnel plots. The review of Storebø and colleagues identified potential bias on their funnel plot because of asymmetry but found no evidence of significant publication bias. Egger’s regression intercept was −0.2260 (two-tailed, p=0.81) [1].

Methylphenidate has been used globally for more than 50 years for treatment of children with ADHD [1]. Methylphenidate’s precise neurochemical mechanism of action is under debate, but it is potentially related to dopaminergic and noradrenergic neurotransmissions in the brain [2]. Methylphenidate can be delivered via immediate, sustained, or extended release. Furthermore, it can be delivered by oral or transdermal routes [1]. The dosage of methylphenidate can vary significantly. The dose can range from 5 mg to 60 mg of methylphenidate per day, and usually about 1.4 mg/kg daily administered in two or three doses [1].

Research waste and late publishing of randomised clinical trials are areas that have received increased focus as of late [3]. A recent study of German University Medical Centers found that prospective registration of interventional trials, within all therapeutic areas, had increased from 33% to 75% between 2009 and 2017 in ClinicalTrials.gov. However, only 41% of included trials had published their result within 2 years of trial completion and 31% had not published their result even after 5 years of trial completion [3].

The German study used 24 months after registration of the protocol as their trial completion date. The European Medical Agency has, however, required sponsors of clinical trials including children to make tabular summary results publicly available within 6 months of a trials primary completion date [4]. Accordingly, we will use this 6-month regulation for considering whether a trial has been published timely or not.

## Objectives

Our primary objective is to assess how many randomised clinical trials of methylphenidate versus placebo or no intervention with or without co-intervention for children and adolescents with ADHD are registered in protocol databases, but never published in the academic literature or as tabular summary results. We will also determine how many trials are published after 2005 without the protocol being registered. The number of participants included in these trials is a secondary objective. Our tertiary objective is to examine the relationship between trial results (positive compared to neutral or negative or to unknown) and the chance of being published. Our fourth objective is to assess the time from registry to publication of randomised clinical trials of methylphenidate versus placebo or no intervention for children and adolescents with ADHD in either a journal or as summary results, and the number of participants in these trials.

## Research questions

How many randomised clinical trials on methylphenidate versus placebo or no intervention with or without co-intervention for children and adolescents with ADHD are registered as protocols of completed and terminated trials in clinical trial registers, but never published as articles in the academic literature? How many trials are published after 2005 without the protocol being registered? And how many participants are used in these trials? What is the relationship between trial result (positive compared to neutral or negative or to unknown) and the chance of being published? To what extent do the trials registered in clinical trial registers follow the 6 month guideline for publication of tabular summary results after the primary completion date? And what are the number of participants in these trials?

## Methodology

### 1. Cohort selection

Method for literature search and validation of findings was inspired by Bruckner and Grzegorzek [5].

A search for trial registers:

Clinicaltrials.org and the EUCTR databases and WHO International Clinical Trials Registry Platform for randomised clinical trials on methylphenidate for ADHD is made with the following inclusion criteria:

- Methylphenidate versus placebo or no intervention with or without co-interventions for children or adolescents with ADHD
- Randomised clinical trial
- Any dosage of methylphenidate
- Any delivery method
- At least 75% children and adolescents with ages less than 18 years.
- Registered between 1999-2022

We will search ClinicalTrials.gov and EUCTR.

The search string used for ClinicalTrials.gov is the following:

Advanced search. Condition or disease: ADHD. Other terms: Methylphenidate. Study type: Intervention study. Age group: 0 to 18 years. Date range: 1999-2022.

The search string used for EUCTR was the following:

Advanced search. Search ADHD AND methylphenidate. Choose under 18 years. Date range: 1999-2022.

After we have found our cohort, Storebø et al. 2023[1] will be used during screening to see if a trial publication was found in the previous data collection, either as included or excluded. Screening will be performed by two independent researchers reaching a consensus on inclusion or exclusion.

### 2. First literature search

Of the included cohort, the following steps will be taken:

- If included in Storebø et al. 2023 [1], data will be extracted via that reference
- Scan for tabular summary results
- Scan for publications uploaded by sponsor/investigator or automatically indexed by registry
- Enter clinical trial identifier in Google Scholar and search first 2 pages for potential match by title.
- Enter clinical trial identifier in Google Scholar and search first 2 pages for potential match by putting the principal investigator name in “quotations”.

If, at any step, a publication is found, it will be verified that it is a results publication and not just another registry protocol or other irrelevant document. Judgement will be made via title and abstract. Full text only if needed. The date and link to publication will be extracted.

If the trial publication does not include the result of the primary outcome, then the search will continue. Grey literature such as posters or presentation slides will be extracted separately in a spreadsheet.

### 3. Validation of findings

An e-mail will be sent to the company or institution that sponsored a trial where no results could be found, and invite the sponsor to either send any relevant publications that were missed or provide a short on-the-record statement on their clinical reporting policies and plans for inclusion in a supplementary annex of the final manuscript. One reminder e-mail will be sent if no response. E-mails will disclose that the study in question is to be used in a study on research waste.

### 4. Second literature search

A second search will be made for published clinical trials where first search returned unsuccessful and the sponsor did not provide the status of the trial

- First, a repeat of the first steps will be made
- Then, PubMed will be searched with the clinical trial identifier
- PubMed will be searched for +”intervention name” and +”condition name”. The first 2 pages will be searched
- Google Scholar will be searched for +”intervention name” and +”condition name”. The first 2 pages will be searched

## Outcomes

The first outcome is the extent of publications of randomised clinical trials of methylphenidate versus placebo or no intervention with or without co-interventions for children or adolescents with ADHD in a peer-reviewed journal or PhD thesis for trials missing summary results in clinical trial databases.

The second outcome is the number of participants in the unpublished trials, the timely published trials and the untimely published trials.

The third oucome is the number of participants in the published trials with and without a relevant registration in the searched registers.

The fourth outcome is the result of a chi-squared test where the chance of being published is compared to positive, neutral, negative and unknown test results.

The fifth outcome is the speed of publication for randomised clinical trials of methylphenidate versus placebo or no intervention with or without co-interventions for children or adolescents with ADHD in a peer-reviewed journal or PhD thesis for trials registered in clinical trial databases. The speed is calculated by the time difference between a trial’s primary completions date and the first published date of the trial in a peer-reviewed journal and/or a registry.

## Summary statistics

Four tables will be made regarding publication speed and research waste as a % of trials, % of patients, # of patients and # of trials. Proportions will be compared with chi-square tests.

**Table 1:** Publication speed and research waste of randomised clinical trials on methylphenidate for children and adolescent with ADHD, % of trials

|  | Timely publication | Delayed publication |  |  | Research waste | Publication speed (average months) |
| --- | --- | --- | --- | --- | --- | --- |
|  | Within 6 months | Within 6 months to 2 years | Within 2 to 3 years | After 3 years | No results |  |
| All trials |  |  |  |  |  |  |

**Table 2:** Publication speed and research waste of randomised clinical trials of methylphenidate in children and adolescent with ADHD, # of trials

|  | Trials total | Timely publication | Delayed publication |  |  | Research Waste |
| --- | --- | --- | --- | --- | --- | --- |
|  |  | Within 6 months | Within 6 months to 2 years | Within 2 to 3 years | After 3 years | No results |
| All trials |  |  |  |  |  |  |

**Table 3:** Publication speed and research waste of randomised clinical trials on methylphenidate for children and adolescent with ADHD, % of patients

|  | % of patients | Timely publication | Delayed publication |  |  | Research waste |
| --- | --- | --- | --- | --- | --- | --- |
|  |  | Within 6 months | Within 6 months to 2 years | Within 2 to 3 years | After 3 years | No results |
| All trials |  |  |  |  |  |  |

**Table 4:** Publication speed and research waste of randomised clinical trials on methylphenidate for children and adolescent with ADHD, # of patients

|  | Patients total | Timely publication | Delayed publication |  |  | Research waste |
| --- | --- | --- | --- | --- | --- | --- |
|  |  | Within 6 months | Within 6 months to 2 years | Within 2 to 3 years | After 3 years | No results |
| All trials |  |  |  |  |  |  |

The output of a chi-squared test will be included, showing the chance of being published compared to positive, neutral, negative and unknown test results.

A Kaplan-Meier curve tracking publication speed will also be included.

## Discussion

A limitation of our study is the use of only Clinicaltrials.org and the EUCTR databases and WHO International Clinical *Trials Registry* Platform for the literature search. This narrows the search somewhat towards western studies, and a more comprehensive list of registers could have been used. This was not possible due to time constraints as well as limited resources.

Another limitation of our study is that we will not be looking at clinical study reports for publication bias. This is also due to time constraints as well as limited resources.

## Data Availability

All data produced are available online at clinicaltrails.gov and the EU clinical trials register

https://www.clinicaltrialsregister.eu/ctr-search/search

